# SARS-CoV-2-specific Humoral Immune Responses after A Single dose of Inactivated Whole-virus SARS-CoV-2 Vaccine in Kidney Transplant Recipients: An Initial Report

**DOI:** 10.1101/2021.07.19.21260694

**Authors:** Jackrapong Bruminhent, Sarinya Boongird, Montira Assanatham, Sasisopin Kiertiburanakul, Kumthorn Malathum, Arkom Nongnuch, Angsana Phuphuakrat, Pongsathon Chaumdee, Sopon Jirasiritham, Chitimaporn Janphram, Sansanee Tossiri, Supparat Upama, Chavachol Sethaudom

**Author notes:** **Correspondence** Chavachol Sethaudom.

## Abstract

We presented an initial pilot study report focused on immunogenicity and safety following an inactivated whole-virus severe acute respiratory syndrome coronavirus 2 vaccination among kidney transplant (KT) recipients. At four weeks after the first dose of vaccine, the level of anti–receptor-binding domain IgG antibody was not significantly different compared to before vaccination in 30 KT recipients (p = 0.45). Moreover, a significant lower mean (95% CI) anti–receptor-binding domain IgG antibody was observed compared to 30 immunocompetent controls (2.4 [95% CI 1.3-3.5] vs. 173.1 [95% CI 88.3-2,457.9] AU/mL, p < 0.001). Mild adverse events included fever (17%) and localized pain at the injection site (14%) were observed after vaccination.

## Brief report

Solid-organ transplant (SOT) recipients are at greater risk of significant coronavirus disease 2019 (COVID-19) related morbidity and mortality.^1^ Immunization against severe acute respiratory syndrome coronavirus 2 (SARS-CoV-2) is considered one of the most promising strategies to alleviate unfavorable consequences among SOT recipients and their allograft.

Several platforms of the SARS-CoV-2 vaccine were developed and utilized variably worldwide among immunocompetent individuals. Data on SARS-CoV-2-specific immune responses among patients with immunocompromised states such as those receiving SOT has been scarce. Few recent studies evaluated the immunogenicity of SARS-CoV-2 messenger RNA vaccine in SOT recipients receiving immunosuppressant revealed an acceptable safety profile. However, a significant proportion of recipients who developed suboptimal immunologic response after a single and double dose of the vaccine is a concern (17% and 54%, respectively).^2,3,4^ On the other hand, immunogenicity, and safety following immunization with an inactivated whole-virus SARS-CoV-2 vaccine among SOT recipients have not been assessed.^5^

A pilot, multi-center, prospective study of adult Asian KT recipients who received a 4-week interval immunization of an inactivated whole-virus SARS-CoV-2 vaccine, CoronaVac^®^ (Sinovac Biotech Ltd.) was conducted from April 2021 to June 2021. SARS-CoV-2-specific humoral immunity was measured before, one month after the first dose, using a quantitative assay of SARS-CoV-2 immunoglobulin G (IgG) assay (Abbott diagnostics) that tests for antibodies against the receptor-binding domain (RBD) of the SARS-CoV-2 spike protein, which reported in arbitrary units (AU)/mL. Those were also referenced to immunocompetent patients. The study was approved by the Institutional Review Board of the Faculty of Medicine Ramathibodi Hospital, Mahidol University, Bangkok, Thailand (MURA2021/242). All patients provided written informed consent prior to participation.

A total of adult 60 patients was recruited into the study and comprised of immunocompetent patients (n=30) and KT recipients (n=30). Among KT recipients, the median age was 47 years (interquartile range [IQR], 41-52 years), and 67% of them were men. The median time since transplant was 3.6 years (IQR, 2-9.5 years). The maintenance immunosuppression regimen included tacrolimus (57%), cyclosporine (29%), corticosteroids (97%), mycophenolic acid (91%), sirolimus (3%), and everolimus (3%). At four weeks after the first dose of vaccine, a mean (95% confidence interval [CI]) of anti–receptor-binding domain IgG antibody was not significantly different compared to before vaccination in all participants (p = 0.45) **Figure 1**. KT recipients developed significantly lower anti–receptor-binding domain IgG antibody compared to immunocompetent patients (2.4 [95% CI 1.3-3.5] vs. 173.1 [95% CI 88.3-2,457.9] AU/mL, p < 0.001). Approximately half of the participants did not report any adverse events (AEs) followed by mild AEs such as fever (17%) and pain at the injection site (14%). No serious AEs observed.

**Figure 1.**
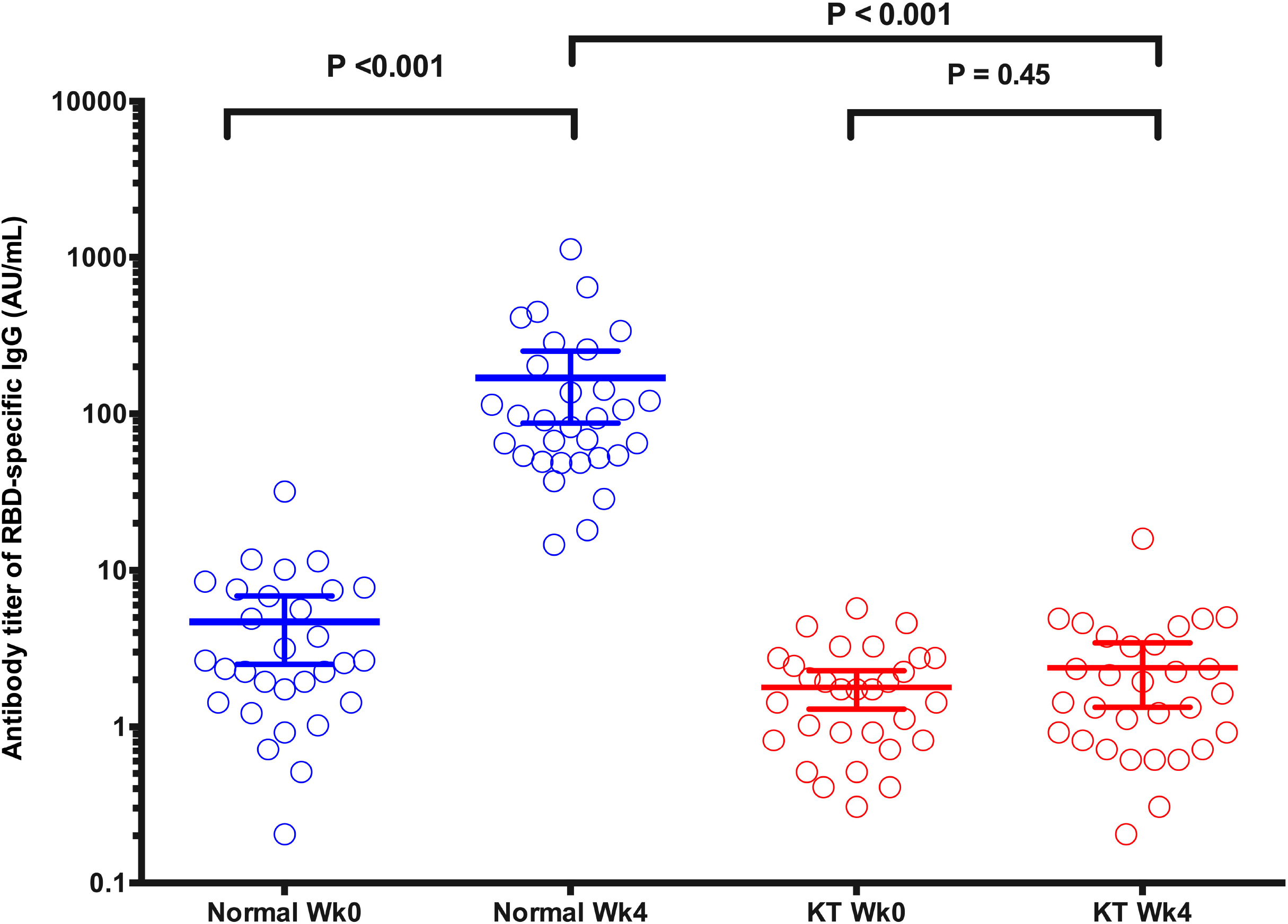
A mean with 95% confidence interval of anti–receptor-binding domain immunoglobulin G antibody in immunocompetent patients and kidney transplant recipients before and at four weeks after a single dose of inactivated whole-virus SARS-CoV-2 vaccine.

Our initial report revealed that KT recipients receiving the first dose of an inactivated whole-virus SARS-CoV-2 vaccine could remain vulnerable to infection, however, with infrequent and mild adverse reactions. A complete result is ongoing, and further large-scale studies reaffirmed this result is needed (Thai Clinical Trials Registry, TCTR20210226002).

## Data Availability

The availability of all data referred to in the manuscript is available upon request.

## Disclosure

The authors of this manuscript have no conflicts of interest to disclose

## Funding

This study was granted by the National Research Council of Thailand (102912).

## Notes

### Competing Interest Statement

The authors have declared no competing interest.

### Clinical Trial

Thai Clinical Trials Registry, TCTR20210226002

### Funding Statement

This study received a grant from the National Research Council of Thailand (NRCT), Ministry of Higher Education, Science, Research, and Innovation of Thailand in collaboration with the Department of Medical Services, Ministry of Public Health of Thailand (102912).

### Author Declarations

The Institutional Review Board of the Faculty of Medicine reviewed and approved the study protocol at Ramathibodi Hospital, Mahidol University, Bangkok, Thailand (approval number: MURA2021/242).

